# Causal meditation analysis to understand how different components of a complex psychological intervention improved symptoms (or not) of depression in Goa, India

**DOI:** 10.1101/2021.10.04.21264478

**Authors:** Nadine Seward, Stijn Vansteelandt, Darío Moreno-Agostino, Vikram Patel, Ricardo Araya

## Abstract

**Background:** Understanding how and under what circumstances a highly effective psychological intervention, improved symptoms of depression is important to bring this evidence-informed intervention to scale, particularly in resource-poor settings. We aim to estimate the indirect effects of potentially important mediators to improve symptoms of depression in the Healthy Activity Program (HAP) trial.

**Methods:** Interventional in(direct) effects were used to simultaneously decompose the total effect of the intervention on depression symptoms measured through the Patient Health Questionnaire (PHQ-9). The following indirect effects were considered: characteristics of sessions including the number of sessions and homework completed; behavioural activation according to an adapted version of the Behavioural Activation for Depression Short Form (BADS-SF), and extra sessions offered to participants who did not respond to the intervention.

**Results:** Of the total effect of the intervention measured through the difference in PHQ-9 scores between treatment arms (mean difference: -2.2, 95% bias-corrected CI: -3.2, -0.8), 45% was mediated through improved levels of behavioural activation (−1.0, -1.3, -0.6). There was no evidence to support the mediating role of characteristics of the sessions nor the extra sessions offered to participants who did not respond to the treatment.

**Conclusions:** Findings from our robust mediation analyses, confirmed the importance of behavioural activation in improving depression symptoms. Contrary to published literature, our findings suggest that neither the number of sessions nor proportion of homework completed, improved outcomes. Moreover, results indicate that the extra sessions were insufficient to improve symptoms of depression for participants who did not respond to the intervention.

## Introduction

Depression is a common mental health disorder that affects an estimated 300 million people worldwide and is the leading mental health cause to the global burden of disease.(Patel V et al., 2016) Depression is also associated with excess mortality and morbidity as well as profound social and economic consequences. Despite this, only 10% or fewer people with depression in low- and middle-income countries (LMICs) have access to effective treatment.

Psychological treatments are the recommended first-line intervention for depression according to the WHO’s Mental Health Gap Action Programme (mhGAP) as they have been shown to be just as effective as pharmacological interventions and have a sustained effect over time.(Who, 2008) However, important barriers to accessing these treatments in LMICs exist, in particular the lack of trained professionals.(Patel V, 2000)

Behavioural activation for depression is particularly applicable to LMICs as lay health workers with limited training can effectively deliver the intervention.(Jacobson N, Martell C, & Dimidjian S, 2001) Behavioural activation improves symptoms of depression mainly by the mechanism of increasing the number of enjoyable or meaningful activities. There is good but not undisputed evidence to support the effectiveness of behavioural activation for depression in LMICs (Araya R et al., 2021; Arjadi et al., 2018; Patel V et al., 2017; Sikander S et al., 2019).

Reasons to explain these differences in effectiveness from different trials may be related to the contexts where the interventions are delivered (middle vs low-income country or urban vs rural), the conditions studied (perinatal depression or comorbid hypertension and depression), mode of delivery (mHealth vs task sharing) and other underlying factors that are often not accounted for.

Characteristics of the sessions are known to influence depression outcomes through different pathways. As an example, evidence suggests that the number of sessions is an important predictor of recovery, with five to six sessions required as a minimum to improve depression outcomes.(Dunn G et al., 2012; Patel V et al., 2017) Analyses that account for the multiple and often interacting pathways through which an intervention operates including both characteristics of the sessions and mechanisms such as behavioural activation will help to determine how to optimise different components in preparation for future scale-up. Comparing findings across different studies will provide insight into what works for whom and under what circumstances.

### The Healthy Activity Programme (HAP)

One of the most successful trials of a lay counsellor-delivered psychological intervention for depression from a LMIC is the HAP trial. (Patel V et al., 2017) HAP is an adapted form of behavioural activation delivered by lay counsellors on a face-to-face modality for participants with moderate to severe depression in primary care settings in Goa, India. The effectiveness of the HAP intervention on remission from depression was evaluated using a randomised controlled trial. Remission from depression was defined by a Patient Health Questionnaire 9 (PHQ-9) score less than 10. Findings suggest that participants in the experimental arm had higher odds in remission from depression compared to the control arm, (adjusted odds ratio 1.61, 95% CI 1.34, 1.93) 3 months after the trial started. Results also suggested increased activity levels in the experimental arm, measured through an adapted version of the Behavioural Activation for Depression Score (BADS-SF) (adjusted mean difference 2.2, 95% CI 1.3, 3). A follow-up study conducted at 12 months after the trial started, demonstrated that the of odds of remission from depression was still higher in the experimental arm compared to the control arm (adjusted odds ratio 1.36; 95% CI 1.15, 1.61; p < 0.009).(Weobong B et al., 2017) However, little is known surrounding how HAP improved recovery from depression, other than findings from a mediation analysis that found 58% of the total effect was mediated through improved activation measured using the BAD-SF.(Weobong B et al., 2017)

Given the potential to bring this intervention to scale, it is important to understand how the HAP intervention can be optimised. The interventional (in)direct effects, is the latest in a series of recent advancements to causal mediation analyses that allows for the decomposition of the total effect of a complex intervention into multiple indirect effects that can characterise specific mechanisms and characteristics of the sessions.(Vansteelandt & Daniel, 2017) One advantage of this approach is that it is possible to simultaneously account for multiple mediators, their interactions and non-linearities, allowing for better insight into the mechanism of impact of an intervention.

We therefore estimated interventional (in)direct effects based on data collected both at three and 12 months after the trial started for the following mediators: characteristics of the sessions (i.e. number of sessions and proportion of homework completed), behavioural activation, and extra sessions a participant received if they did not respond to treatment. It is hoped that findings will provide further insights into what components of the intervention worked (or not).

## Methods

### Setting

We used data from a previously conducted randomised controlled trial (HAP) that took place between October 2013 and July 2015 in primary health care centres in Goa, India, that included information on symptoms of depression and important mediators at three and 12 months after the trial started.(Patel V et al., 2017; Weobong B et al., 2017) The original trial was registered with the ISRCTN registry, number ISRCTN95149997.

### Design

HAP was a parallel arm, individually randomised controlled trial with equal allocation of participants between arms. Participants aged between 18-65 years were recruited from 10 primary health centres. Eligibility criteria included a probable diagnosis of moderately to severe depression determined with a PHQ-9 score greater than 14. Pregnant women and participants who needed urgent medical attention or who were unable to communicate were excluded.

### The intervention

The intervention (HAP) was a manualised psychological intervention based on behavioural activation for depression that primarily involved strategies to increase the number of enjoyable activities a person engaged with.(Patel V et al., 2017; Patel et al., 2014) Other strategies were also included after exploring their acceptability, appropriateness and feasibility in the local context including need-based strategies that addressed interpersonal triggers, problem-solving, relaxation and enlisting social support tailored to the specific need of the individuals.(Chowdhary N et al., 2016)

The experimental arm received up to eight sessions (described below) that lasted between 30 and 40 minutes at weekly intervals over a 3-month period. The sessions were usually face to face at the Primary Health Centre, or the patient’s home. Telephone sessions were used only when strictly necessary. The intervention was organized in three phases. The first phase (sessions 1-2) was primarily used to engage the participant, establish an effective relationship, explain the objectives of the sessions including behavioural activation, and to elicit a commitment for the HAP intervention. The middle phase (sessions 3-4) assessed activation targets and encouraged activation, identifying barriers to activation, and learning to overcome these and how to solve or cope with life problems. The ending phase (sessions 5-6) reviewed and strengthened gains the patient made during treatment in order to prevent relapse. If a participant did not respond to treatment by the third or fourth session, two additional middle-phase sessions were offered resulting in these patients attending a total of seven to eight sessions.

HAP was delivered by lay counsellors who had completed at least the 10^th^ grade of education and were fluent in local languages. Counsellors were also required to meet pre-defined competency standards.(Singla D et al., 2014) Training took place over a three-week period. Counsellors received weekly peer-led supervision in groups and individual supervision twice a month.

Enhanced usual care (EUC) was offered to participants in both arms of the trial.(Patel V et al., 2017) EUC involved screening results for depression being shared with both patients and physician. Physicians were also trained on how to use a contextualised version of the mhGAP guidelines, including when and where to refer for psychiatric care.

### Measures

#### Exposure

Our exposure of interest was the HAP intervention that was offered to participants in the experimental arm of the trial only.

#### Outcome

The HAP trial had two primary outcomes: remission from depression evaluated using the PHQ-9 (a score of less than 10 was considered as remission from depression) and the total depression score evaluated using the Becks Inventory for Depression version II (BDI-II). Outcomes were evaluated at both 3 months and 12 months after the trial started. For our analysis, we selected the secondary outcome of PHQ-9 score at 12 months as our primary outcome for two reasons: (1) the PHQ-9 had been validated for this study setting whereas the BDI-II had not; (2) the PHQ-9 total score is a discrete outcome that allows a finer grained estimate of the indirect effect of the different mediators, compared to a binary outcome of remission from depression (yes/no). Response options generate a continuous score ranging from 0-27 since each of the nine items can be scored from zero (no symptoms) to three (nearly every day). Scores between ten and 14 represent moderate depression, and scores between 15 and 27 moderately severe to severe depression symptoms.(Kroenke K, Spitzer R, & Williams J, 2001)

#### Mediators

Causal mediation analyses require obtaining estimates in both the exposed participants, as well as the unexposed (i.e. the counterfactual). Where mediators were measured in both the experimental and control arms, estimates from participants in the control arm can be treated as the unexposed. However, where a mediator was only measured in the experimental arm, it was necessary to create a separate category for participants who had not been exposed to the mediator of interest but were still in the experimental arm.

### Therapeutic process indicators (M1)

Characteristics associated with the delivery of the sessions were measured for participants in the experimental arm only and considered as potential mediator of interest as it was possible these factors might influence both behavioural activation levels and depression. The first characteristic that we accounted for is the number of sessions completed that are categorised to reflect the phases of the HAP intervention that a participant completed (M1a: no sessions (unexposed); sessions one and two (phase 1), sessions three and four (phase 2); sessions five through eight (phase 3).

The second characteristic of the sessions that we accounted for is a participant’s self-reported completion of assigned tasks outside of sessions (homework) to improve activity levels. At each session except for the first one, activity monitoring charts were completed indicating whether a participant completed homework outside of the sessions. These self-reported activity charts were scored using the following criteria: completely (scored 2), partially (scored 1), or not at all (scored 0). Based on this variable, we calculated a score representing the proportion of homework completed (M1b: 0= none (unexposed); 1= > 0% and ≤ 50%; 2=> 50%).

### Level of behavioural activation (M2)

Behavioural activation was measured for participants in both the experimental (exposed) and control arms (unexposed) at 3 months after the trial started. An adapted version of the Behavioural Activation for Depression Scale Short Form (BADS-SF) was used to capture activity levels that reflect the level of behavioural activation due to the HAP intervention.(Manos R, Kanter J, & Luo W, 2011) This adapted version was developed for the HAP trial, based on pilot data, and included five of the nine original items: (1) are you content with the amount and types of things you did; (2) did you engage in many different activities; (3) were you an active person and accomplished the goals you set out to do; (4) did you spend long periods thinking over and over about your problems, and (5) did you do things that were enjoyable. Total scores ranged between 0 and 20.

### Extra sessions received in instances of non-response to the intervention (M3)

Adding extra sessions for participants who do not respond to the intervention may help to improve symptoms of depression. Therefore, if a participant did not respond to the intervention by session five, they were offered two additional sessions. Estimating this indirect effect via the additional sessions involves two variables including non-response to treatment (M3a: 0=non response (unexposed); 1=responded to treatment) and the number of extra sessions received (M3b: 0=no extra sessions (unexposed); 1=one extra session; 2=two extra sessions). Appendix 1 details how non-response to the sessions was determined.

#### Mediator-outcome confounders

Due to the randomised nature of the exposure, it was not necessary to account for confounders for the association between the exposure and the outcome, or between the exposure and the mediators. However, it was necessary to account for confounders potentially distorting the association between the mediator and the outcome (mediator-outcome confounders). We considered all demographic characteristics as potential mediator–outcome confounders. The selection process for these confounders is described in the section on statistical methods.

### Statistical methods

#### General

To better understand the relationship between different mediators and depression outcomes, we compared characteristics of the sessions (i.e. M1a - number of sessions and M1b proportion of homework completed), behavioural activation (M2), non-response to the intervention (M3a), and the number of extra sessions attended for participants who did not respond to HAP (M3b), with the outcome remission from depression (determined by a PHQ-9 score less than 10) for participants in the experimental arm only. Differences in baseline characteristics between treatment arms can be found in previous publications.(Patel V et al., 2017; Weobong B et al., 2017)

#### Mediation analysis

We aimed to investigate the extent to which symptoms of depression measured at 12 months using the PHQ-9 questionnaire, were explained by the direct effect and indirect effects of the intervention (Figure 1). To achieve these objectives, we used the interventional (in)direct effects approach to mediation analysis to understand population level effects relevant to this analysis.(Vansteelandt & Daniel, 2017) Findings for this analyses are reported according to guidelines for reporting mediation analyses (AGReMA statement).(Lee H et al., 2021)

**Figure I:**
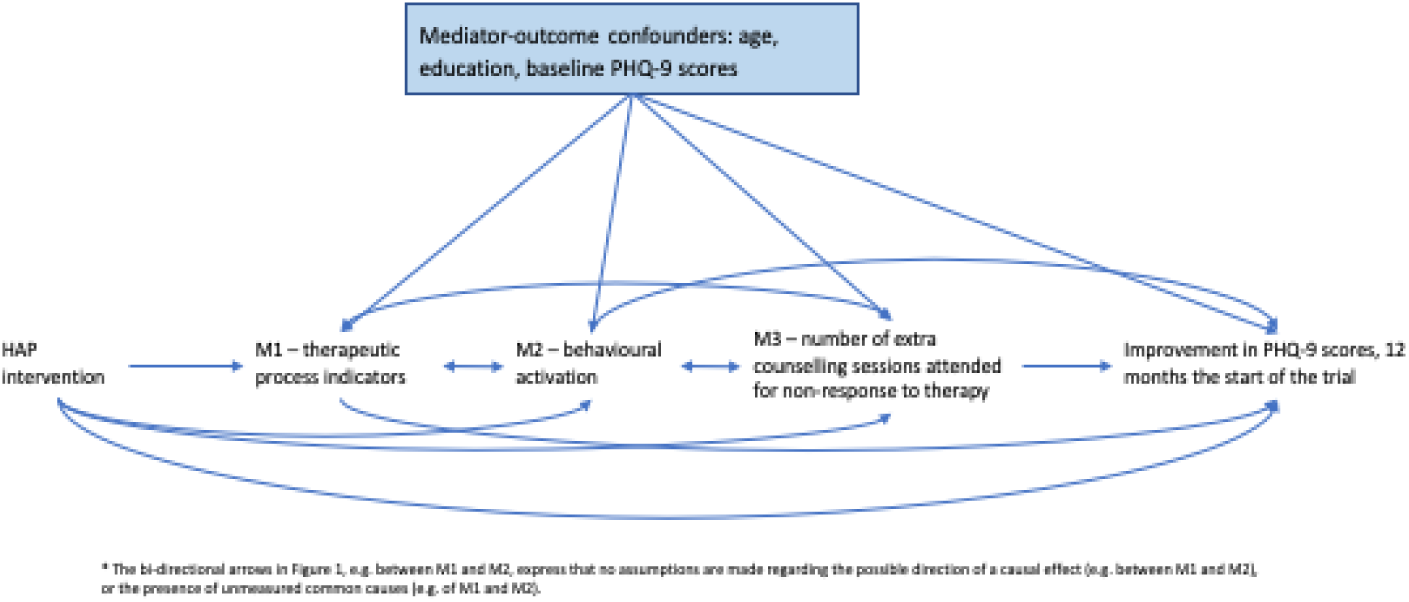
Causal model demonstrating the proposed pathways through which the HAP intervention may improve remission from depression.

#### Decomposition of total effect of the HAP intervention into direct and indirect effects

The first step of the mediation analyses involved decomposing the total effect of the HAP intervention, into path-specific indirect effects and the direct effect. In order for the decomposition to be valid, the sum of the different path specific effects including the direct effect, through which the effects of the intervention is mediated, must be the same as the total effect of the intervention.(Daniel R, De Stavola B, Cousens S, & Vansteelandt S, 2015)

With three mediators, the following decompositions are required to sum the total effect: an estimate that does not involve any mediators (level 0 - all mediators set to unexposed); an estimate that includes one mediator (level 1 – one mediator exposed, the rest unexposed); two mediators (level 2 – two mediators set to exposed, one to unexposed), and three mediators (level 3 – all three mediators set to exposed).

Setting a mediator at an exposed status indicates the pathway includes the effect of this mediator. Likewise, setting a mediator to an unexposed status, indicates the pathway excludes the effect of this mediator (i.e. setting M2 and M3 to unexposed indicted the pathway includes M1 only). Table 1 describes the total effect and decompositions for the direct and indirect effects of the HAP intervention that sum the total effect.

**Table I:** Decompositions for the direct and indirect effects of the HAP intervention that sum the total effect.

| Estimate | Description | M1 | M2 | M3 |
| --- | --- | --- | --- | --- |
| Total effect | Difference between predicted PHQ-9 scores between the experimental and control arm | NA | NA | NA |
| Direct effect | Difference between predicted PHQ-9 scores in the intervention and control arms, whilst fixing mediators to unexposed status (level 0) | unexposed | unexposed | unexposed |
| M1 – characteristics of the sessions | Difference between predicted PHQ-9 scores when changing M1 from levels in the exposed, to levels in the unexposed, in the experimental arm (level one) | exposed to unexposed | unexposed | unexposed |
| M2 – levels of behavioural activation | Difference between predicted PHQ-9 scores when changing M2 from levels in the exposed, to levels in the unexposed, in the experimental arm (level 2) | exposed | exposed to unexposed | unexposed |
| M3 – additional sessions offered in instances of non-response | Difference between predicted PHQ-9 scores when changing M3 from levels in the unexposed, to levels in the exposed, in the experimental arm (level | exposed | exposed | exposed to unexposed |
|  | 3) |  |  |  |

### Estimation

Estimation for the different effects was based on Monte Carlo integration using 1,000 fold expanded dataset. Estimates for the total, direct, and indirect effects were obtained by running regression models for the outcome of PHQ-9 score, separately in the exposed and unexposed, whilst setting the mediators at a random subject-specific distribution (Table 1). Further details of the estimation methods can be found in Appendix 1.

### Model fit

Regression models used to estimate the total, direct and indirect effects, included any mediator-outcome confounder that improved model fit as indicated by the Akaike Information Criterion (AIC).(Burnham K & Anderson D, 2004) Results indicate that age, education, and baseline PHQ-9 scores are important confounders. These models also included mediators set to relevant exposed/unexposed status (Table 1).

The regression models used to set the mediators at random, subject-specific draws for the unexposed/exposed status, included any variable that improved model fit as indicated by the Akaike Information Criterion (AIC),(Burnham K & Anderson D, 2004) but not known to be potentially influenced by them (i.e., thus excluding outcome, and other mediators for instance). This ensured that mediator values drawn were more specific to the considered individual, thereby providing better insight into mechanism. Models for the different mediators used a combination of predictors including age, education, baseline PHQ-9 scores, participants expectations of treatment, and marital status. Any relevant non-linearities and interactions were included in all models if determined to be significant at the five percent level, using the post-estimation *testparm* command in Stata.

### Assumptions

The interventional effects have important underlying assumptions that will influence the validity of our findings if violated. Reassuringly, due to the randomised nature of the HAP trial, many of the assumptions with the interventional effects are fulfilled. The interventional effects capture the components of the total effect mediated by the different mediators, even when the structural dependence between multiple mediators is unknown (i.e. direction of the causal effects between the multiple mediators is unknown, or if there is unmeasured common causes of the mediators (Figure 1)). The main assumption relevant to our study is that there are no unmeasured mediator-outcome confounders.

### Missing data

There were missing data for the BADS-SF variable (measuring M2) at 3 months (n=28, 5.7%) and the PHQ-9 variable at 12 months (n=47, 9.3%). To account for this, we implemented single stochastic imputation using chained equations with 10 burn-in iterations, under the assumption that data was missing at random (MAR). Details of the missing data analysis can be found in Appendix 2.

### Ethical approval and consent

The trial protocol received ethical approval from the Sangath and LSHTM Institutional Review Boards.(Patel et al., 2014) Written or witnessed verbal (if the participant is illiterate) informed consent was mandatory for enrolment. All consent procedures were audio-taped, with the patient’s approval, for quality assurance.

## Results

There were 493 participants included in the study, with 248 (50%) allocated to the control arm with EUC alone, and 245 (50%) to the experimental arm with EUC plus HAP. Table 2 compares mediators measured at 3 months, between participants with remission from depression (PHQ-9 <10) and participants with depression (PHQ-9 >=10) measured at 12 months after the trial started, in the experimental arm only. The mediator representing characteristics of the counselling sessions (M1) demonstrated a non-linear relationship with remission from depression. As an example, participants who attended 0-2 sessions were more likely to recover from depression compared to participants who attended 6-8 sessions. Participants who attended five sessions were more likely to recover from depression than participants who attended any other number of sessions. Findings suggest at 12 months, participants who had sustained remission from depression, had higher mean behavioural activation levels measured at 3 months (compared to participants who did not recover from depression (M2).

**Table 2.** Comparison of mediators between participants with and without remission from depression at 12 months, in the experimental arm of the trial only.

| Mediators | Remission from depression determined using PHQ-9 scores less than 10 at 12 months in the experimental group only (n=245)<sup>a</sup> |  | Overall |
| --- | --- | --- | --- |
|  | No remission from depression (N=89) (PHQ-9 > 10) N (%) | Remission from depression (N=156) (PHQ-9 < 10) N (%) |  |
| Number of sessions attended |  |  |  |
| 0 | 6 (6.7) | 13 (8.4) | 19 (7.8) |
| 1 | 4 (4.4) | 16 (10.9) | 20 (8.2) |
| 2 | 7 (7.8) | 17 (11.0) | 24 (9.8) |
| 3 | 2 (2.2) | 3 (1.9) | 5 (2.0) |
| 4 | 2 (2.2) | 2 (1.3) | 4 (1.6) |
| 5 | 11 (12.2) | 47 (30.3) | 58 (23.7) |
| 6 | 23 (25.6) | 31 (20.0) | 54 (22.0) |
| 7 | 10 (11.1) | 10 (6.5) | 20 (8.2) |
| 8 | 25 (27.8) | 16 (10.3) | 41 (16.7) |
| **M1a: Number of phases of sessions completed** |  |  |  |
| None (no sessions completed) | 6 (6.7) | 13 (8.4) | 19 (7.8) |
| One (sessions 1 to 2 completed) | 11 (12.2) | 33 (21.3) | 44 (18.0) |
| Two (sessions 3 to 4 completed) | 4 (4.4) | 5 (3.2) | 9 (3.7) |
| Three (sessions 5 to 8 completed) | 69 (76.7) | 104 (61.1) | 173 (70.6) |
| **M1b: Proportion of assigned activities completed** |  |  |  |
| No homework | 13 (14.4) | 34 (21.9) | 47 (19.2) |
| 1 – 50% | 14 (15.6) | 25 (16.1) | 39 (16.0) |
| 50 – 100% | 63 (70.0) | 96 (61.9) | 159 (64.9) |
| **M2: Adapted BADS – SF score (mean, standard deviation) measured at 3 months** | 9.5 (4.8) | 13.4 (4.3) | 12.0 (4.7) |
| **M3a: Participants response to HAP measured by PHQ-9 score greater than 9 at 3<sup>rd</sup> or 4<sup>th</sup> session<sup>b</sup>** |  |  |  |
| Did not respond to HAP | 56 (62.2) | 63 (40.7) | 119 (48.6) |
| Responded to HAP | 34 (37.8) | 92 (59.4) | 126 (51.4) |
| **M3b: Number of extra sessions attended if participant did not respond to HAP** |  |  |  |
| None | 55 (61.1) | 129 (83.2) | 184 (75.1) |
| One | 10 (11.1) | 10 (6.5) | 20 (8.2) |
| Two | 25 (27.8) | 16 (10.3) | 41 (16.7) |

<sup>a</sup> Missing data has been imputed by trial arm, using single imputation stochastic models adjusted for any factors that could potentially influence missingness
<sup>b</sup> Table 1 describes how non-response to counselling was determined

**Table 3:** Total effect and interventional in(direct) effects for the HAP intervention at 12 months.

| Effect | Estimates (bias-corrected 95% CI)<sup>a,b,c,d</sup> |
| --- | --- |
| Total effect | -2.2 (-3.3 to -0.8) |
| Interventional direct effect | -3.3 (-5.6 to -0.5) |
| Interventional indirect effect through sessions only (M1) | 2.9 (-1.3 to 5.0) |
| Interventional indirect effect through BA levels only (M2) | -1.0 (-1.3 to -0.6) |
| Interventional indirect effect through extra sessions | -0.8 (-2.1 to 1.2) |
| Interventional indirect effect through the dependence of BA levels on sessions (M3) | 0.0 (-0.6 to 0.7) |

<sup>a</sup> Estimates have been adjusted for mediator-outcome confounders of baseline PHQ-9 scores, age and education.
<sup>b</sup> Estimation for the different effects was based on Monte Carlo integration using 1,000 fold expanded dataset
<sup>c</sup> Bias-corrected confidence intervals were based on nonparametric bootstrap with 1000 resamples
<sup>d</sup> Missing data has been imputed by trial arm, using single imputation stochastic models adjusted for any factors that could potentially influence missingness

### Mediation

Table three demonstrates that at 12 months, the total mean difference in PHQ-9 scores between the experimental and control arms (adjusted mean difference in PHQ-9 scores: -2.2, bias corrected 95% CI: -3.3, -0.8), 45% was mediated through indirect effects via activity levels measured using the adapted version of the BADS-SF (adjusted mean difference in PHQ-9 scores attributable to pathways via BADS-SF -1.0, -1.3, -0.6). There was no evidence to support mediation through indirect effects via characteristics of the sessions including number of sessions and proportion of homework completed (2.9, -1.3, 5.0). The additional sessions offered to non-responders to the intervention did not improve depression scores either (−0.8, -2.1, 1.2). Findings also suggest that the HAP intervention improves symptoms of depression through mechanisms other than those explained with our mediators (−3.3, -5.6, -0.5).

## Discussion

Applying robust, interventional (in)direct effects has allowed us to understand what components of the HAP intervention are effective, and what other components may need to be revised and/or modified before considering scale-up. Specifically, the indirect effect mediated through behavioural activation significantly improved symptoms of depression. However, the indirect effect mediated through both the characteristics of the sessions and the extra sessions offered to participants who did not respond to the intervention, did not appear to improve symptoms of depression.

The main strategy included in the HAP intervention was behavioural activation for depression.(Chowdhary N et al., 2016) Investigators theorised that structuring the sessions to encourage participants to engage with personally chosen enjoyable or meaningful activities outside of the sessions, would improve symptoms of depression. The results from our analysis support this approach, whereby levels of behavioural activation captured using the adapted BADS-SF, improved symptoms of depression. Our findings were also supported by a Cochrane systematic review that found behavioural activation may be more effective than medication in improving symptoms of depression.(Uphoff E et al., 2020) Results from other mediation analyses evaluating the role of behavioural activation are mixed.(Janssen Np et al., 2021) Where some mediation analyses support our findings(Seeley J et al., 2019; Singla D et al., 2021; Weobong B et al., 2017), others show the opposite.(Richards D et al., 2016) However, comparisons are difficult given the different contexts, population under investigation, study designs, conditions being treated, interventions provided, and methods used to conduct the analyses.

Our estimates suggest that characteristics of the sessions we had available, including the number of phases of the intervention the participant attended and the proportion of homework completed, did not influence depression outcomes. Results from the univariate analysis can help to explain this issue. Participants were less likely to recover from depression if they completed more than five sessions, compared to participants who completed five sessions. Likewise, participants were less likely to recover from depression, if they completed more than 50% of their homework, compared to participants who completed less homework. A likely explanation for this phenomenon is that participants who received a more intense intervention (i.e. attended more than five sessions and completed more than 50% of the assigned homework) had more severe forms of depression and might have needed to continue therapy because of non-response. To address this matter, we created an additional mediator to estimate the role of the extra sessions attended by participants who did not respond to the intervention, in improving symptoms of depression. Our findings did not indicate any benefit to receiving the extra sessions suggesting that if the HAP intervention is brought to scale, it will be important to bear this in mind when deciding on how to improve symptoms of depression among non-responders.

Different theoretical approaches to implementation research, other than those driven by a positivist, deductive approach may help to address these findings. As an example, a realist evaluation can be used to better understand why some participants did not respond to the intervention, and under what circumstances.(Pawson & Tilley, 1997) Implementation science theories and frameworks can also be used to help inform why the extra sessions did not improve symptoms of depression.(Nilsen P, 2015) Findings from such investigations could then be used to modify the intervention and future trials explore the effectiveness of the adaptations.

Estimates for our direct effect, indicates that a large proportion of the total effect is still unknown. These results are key, as they suggest that there were characteristics of the HAP intervention not captured with the mediators that we had available, that helped to improve symptoms of depression. Indeed, there were four domains of strategies included as part of the sessions: engagement (psychoeducation, family psychoeducation and treatment planning); behavioural activation; need-based strategies (i.e. addressing interpersonal triggers, problem-solving, relaxation), and social integration.(Chowdhary N et al., 2016) It is entirely conceivable that the effect of the domains not captured with the mediators we had available, was expressed through the direct effect. As an example, HAP investigators theorised that the need-based strategies that were used to address problem-solving, relaxation and enlisting social support, would improve life context, reduce life problems, and eventually alleviate symptoms of depression. This issue highlights the importance of identifying and subsequently collecting relevant data on potential mediators when planning studies. This could provide greater insights into how the intervention can be optimised for future scale-up.

### Limitations

When applying the interventional effects to a randomised trial, the main underlying assumption is that all important mediator-outcome confounders are accounted for. Failing to do so, can potentially bias all estimates including the direct and indirect effects. As an example, non-compliers are often quite different from compliers in terms of depression that may distort the relationships between the direct and indirect effects. Carefully planning a trial a priori can help to ensure any important mediator-outcome confounders are accounted for, therefore reducing bias.(Vo T, Superchi C, Boutron I, & Vansteelandt S, 2020)

Ensuring the variables that capture the mediators are reliable and valid are essential to ensure estimates from meditation analysis are unbiased. Although our outcome measure PHQ-9 had been validated for in this setting, this was not the case with the adapted version of the BAD-SF. To ensure unbiased estimates, future trials should ensure that data collection instruments that are adapted to the trial setting have psychometric properties tested a priori.

Findings of our mediation analyses need to be interpreted with caution when generalising to different contexts. Although these results are relevant for scaling-up the HAP intervention in a similar context to where the trial was originally conducted. By conducting robust mediation analyses for similar interventions in different context, we will be able to better understand what components of the intervention worked for whom and under what circumstances.

## Conclusions

This paper uses a robust approach to mediation analysis to understand how complex psychological therapies work and under what circumstances, something that can potentially be used to inform policy.(Vanderweele T, 2013; Vansteelandt & Daniel, 2017) Not only do our findings reinforce the role of behavioural activation found with other mediation analyses,(Singla D et al., 2021; Weobong B et al., 2017) but we now understand in the context of the HAP trial, that the number of sessions attended and the amount of homework completed is not necessary indicative of an improvement in symptoms of depression symptoms. Moreover, estimates suggest that attending an additional one to two sessions if participants do not respond to the intervention did not improve symptoms of depression. More is needed to understand how interventions such as HAP, can be adapted, in order to improve outcomes in the non-responders.

## Data Availability

data not publicaly available

## Abbreviations

LIMCs: Low- and middle-income countries
WHO: World Health Organization
mhGAP: WHO’s Mental Health Gap Action Programme
HAP: The Healthy Activity Programme
BADS-SF: Behavioural Activation for Depression Score
PHQ-9: Patient Health Questionnaire 9
(EUC): Enhanced usual care
(BDI-II): Becks Inventory for Depression version II

## Acknowledgments

We would like to acknowledge the participants in the trial and the authors who wrote the protocol paper and subsequent papers for the intervention development and trial findings. We would also like to acknowledge the Wellcome Trust Senior Research Fellowship grant to VP (091834) to carry out the original work.

## End materials

### Funding statement

This research received no specific grant from any funding agency, commercial or not-for-profit

### Conflicts of Interest

None

### Ethical standards

The authors assert that all procedures contributing to this work comply with the ethical standards of the relevant national and institutional committees on human experimentation and with the Helsinki Declaration of 1975, as revised in 2008.” The authors also assert that all procedures contributing to this work comply with the ethical standards of the relevant national and institutional guides on the care and use of laboratory animals.

## Notes

### Competing Interest Statement

The authors have declared no competing interest.

### Funding Statement

no funding was received

### Author Declarations

The trial protocol received ethical approval from the Sangath and LSHTM Institutional Review Boards.(Patel et al., 2014) Written or witnessed verbal (if the participant is illiterate) informed consent was mandatory for enrolment.

## References

Araya R, Menezes PR, Claro HC, Brandt, L. R., Daley, K. L., Quayle, J., … Miranda, J. J. (2021). Effect of a Digital Intervention on Depressive Symptoms in Patients With Comorbid Hypertension or Diabetes in Brazil and Peru: Two Randomized Clinical Trials. JAMA, 325(18), 1852–1862. doi:10.1001/jama.2021.4348

Arjadi, R., Nauta, M. H., Scholte, W. F., Hollon, S. D., Chowdhary, N., Suryani, A. O., … Bockting, C. L. H. (2018). Internet-based behavioural activation with lay counsellor support versus online minimal psychoeducation without support for treatment of depression: a randomised controlled trial in Indonesia. Lancet Psychiatry, 5(9), 707–716. doi:10.1016/S2215-0366(18)30223-2

Burnham K, & Anderson D. (2004). Multimodel Inference: Understanding AIC and BIC in Model Selection. Sociological Methods & Research, 33(2), 261–304. doi:10.1177/0049124104268644

Chowdhary N, Anand A, Dimidjian S, Shinde, S., Weobong, B., Balaji, M., … Patel, V. (2016). The Healthy Activity Program lay counsellor delivered treatment for severe depression in India: systematic development and randomised evaluation. The British journal of psychiatry : the journal of mental science, 208(4), 381–388. doi:10.1192/bjp.bp.114.161075

Daniel R, De Stavola B, Cousens S, & Vansteelandt S. (2015). Causal mediation analysis with multiple mediators. Biometrics, 71(1), 1–14. doi:https://doi.org/10.1111/biom.12248

Dunn G, Fowler DRR., Freeman, D., Kuipers, E., Smith, B., … Bebbington, P. (2012). Effective elements of cognitive behaviour therapy for psychosis: results of a novel type of subgroup analysis based on principal stratification. Psychol Med, 42(5), 1057–1068. doi:10.1017/s0033291711001954

Jacobson N, Martell C, & Dimidjian S. (2001). Behavioral activation treatment for depression: Returning to contextual roots [Press release]

Janssen NP, Hendriks GJ, Baranelli CT, Lucassen, P., Oude Voshaar, R., Spijker, J., … Huibers, M. J. H. (2021). How Does Behavioural Activation Work? A Systematic Review of the Evidence on Potential Mediators. Psychotherapy and Psychosomatics, 90(2), 85–93. doi:10.1159/000509820

Kroenke K, Spitzer R, & Williams J. (2001). The PHQ-9: validity of a brief depression severity measure. Journal of general internal medicine, 16(9), 606–613. doi:10.1046/j.1525-1497.2001.016009606.x

Lee H, Cashin A, Lamb S, Hopewell, S., Vansteelandt, S., VanderWeele, T. J., … group, A. (2021). A Guideline for Reporting Mediation Analyses of Randomized Trials and Observational Studies: The AGReMA Statement. JAMA, 326(11), 1045–1056. doi:10.1001/jama.2021.14075

Manos R, Kanter J, & Luo W. (2011). The behavioral activation for depression scale-short form: development and validation. Behav Ther, 42(4), 726–739. doi:10.1016/j.beth.2011.04.004

Nilsen P. (2015). Making sense of implementation theories, models and frameworks. Implementation Science, 10(1), 53. doi:10.1186/s13012-015-0242-0

Patel V. (2000). The need for treatment evidence for common mental disorders in developing countries. Psychol Med, 30(4), 743–746. doi:10.1017/s0033291799002147

Patel V, Chisholm D, Parikh R, Charlson, F. J., Degenhardt, L., Dua, T., … Whiteford, H. (2016). Addressing the burden of mental, neurological, and substance use disorders: key messages from Disease Control Priorities, 3rd edition. Lancet, 387(10028), 1672–1685. doi:10.1016/s0140-6736(15)00390-6

Patel V, Weobong B, Weiss H, Anand, A., Bhat, B., Katti, B., … Fairburn, C. G. (2017). The Healthy Activity Program (HAP), a lay counsellor-delivered brief psychological treatment for severe depression, in primary care in India: a randomised controlled trial. The Lancet, 389(10065), 176–185. doi:10.1016/S0140-6736(16)31589-6

Patel, V., Weobong, B., Nadkarni, A., Weiss, H. A., Anand, A., Naik, S., … Kirkwood, B. (2014). The effectiveness and cost-effectiveness of lay counsellor-delivered psychological treatments for harmful and dependent drinking and moderate to severe depression in primary care in India: PREMIUM study protocol for randomized controlled trials. Trials, 15, 101. doi:10.1186/1745-6215-15-101

Pawson, R., & Tilley, N. (1997). Realistic evaluation. London: Sage.

Richards D, Ekers D, McMillan, D., Taylor, R. S., Byford, S., Warren, F. C., … Finning, K. (2016). Cost and Outcome of Behavioural Activation versus Cognitive Behavioural Therapy for Depression (COBRA): a randomised, controlled, non-inferiority trial. The Lancet, 388(10047), 871–880. doi:10.1016/S0140-6736(16)31140-0

Seeley J, Sheeber L, Feil E, Leve, C., Davis, B., Sorensen, E., … Allan, S. (2019). Mediation analyses of Internet-facilitated cognitive behavioral intervention for maternal depression. Cognitive behaviour therapy, 48(4), 337–352. doi:10.1080/16506073.2018.1513554

Sikander S, Ahmad I, Atif N, Zaidi, A., Vanobberghen, F., Weiss, H. A., … Rahman, A. (2019). Delivering the Thinking Healthy Programme for perinatal depression through volunteer peers: a cluster randomised controlled trial in Pakistan. The Lancet Psychiatry, 6(2), 128–139. doi:10.1016/S2215-0366(18)30467-X

Singla D, MacKinnon D, Fuhr D, Sikander S, Rahman A, & Patel V. (2021). Multiple mediation analysis of the peer-delivered Thinking Healthy Programme for perinatal depression: findings from two parallel, randomised controlled trials. The British Journal of Psychiatry, 218(3), 143–150. doi:10.1192/bjp.2019.184

Singla D, Weobong B, Nadkarni A, Chowdhary, N., Shinde, S., Anand, A., … Patel, V. (2014). Improving the scalability of psychological treatments in developing countries: an evaluation of peer-led therapy quality assessment in Goa, India. Behav Res Ther, 60(100), 53–59. doi:10.1016/j.brat.2014.06.006

Uphoff E, Ekers D, Robertson L, Dawson, S., Sanger, E., South, E., … Churchill, R. (2020). Behavioural activation therapy for depression in adults. Cochrane Database of Systematic Reviews(7). doi:10.1002/14651858.CD013305.pub2

VanderWeele T. (2013). Policy-relevant proportions for direct effects. Epidemiology, 24(1), 175–176. doi:10.1097/EDE.0b013e3182781410

Vansteelandt, S., & Daniel, R. M. (2017). Interventional effects for mediation analysis with multiple mediators. Epidemiology, 28(2), 258–265. doi:10.1097/EDE.0000000000000596

Vo T, Superchi C, Boutron I, & Vansteelandt S. (2020). The conduct and reporting of mediation analysis in recently published randomized controlled trials: results from a methodological systematic review. Journal of Clinical Epidemiology, 117, 78–88. doi:https://doi.org/10.1016/j.jclinepi.2019.10.001

Weobong B, Weiss H, McDaid D, Singla, D. R., Hollon, S. D., Nadkarni, A., … Patel, V. (2017). Sustained effectiveness and cost-effectiveness of the Healthy Activity Programme, a brief psychological treatment for depression delivered by lay counsellors in primary care: 12-month follow-up of a randomised controlled trial. PLoS Medicine, 14(9), e1002385. doi:10.1371/journal.pmed.1002385

WHO. (2008). MhGAP. Mental Health Gap Action Programme. Scaling up care for mental, neurological and substance use disorders. Retrieved from Geneva:

