## Appendix 1 for "Causal meditation analysis to understand how different components of a complex psychological intervention improved symptoms (or not) of depression in Goa, India"

**Appendix 1 – supplementary statistical methods**

Assessing for whether a participant was not responding to treatment and therefore would be offered more sessions.

It is possible, some of the effect of the HAP intervention was mediated through the extra sessions offered to some of the participants if they were not responding to the sessions. Estimating this indirect effect requires an additional mediator, measured using two variables: whether a participant responded to the sessions (M3a), and: the number of extra sessions received (M3b).

*M3a: non-response to sessions*

From the second session onwards, participants were assessed for their response to the intervention using the PHQ-9 questionnaire. According to the HAP program manual, a participant is considered to not be responding and offered additional sessions if they had a PHQ-9 score greater than nine at sessions three or four.(25) Additionally, if a participant scores positive for question 9 on the PHQ-9 questionnaire (question assessing suicide risk), additional sessions were offered. Capturing non-response to the intervention was not straight forward, given not all participants completed the sessions as intended (i.e. dropped out before session five) for reasons that are unknown. Additionally, only the total in-session PHQ-9 scores were available and therefore it was not possible to know if a participant indicated a suicide risk when completing question nine of the questionnaire. Supplementary table 1 describes how non-response to sessions was determined.

**Table 1: Criteria used to determine whether a participant responded to HAP**

| **Number of sessions completed** | **Non-response** | **Response** | **Justification** |
| --- | --- | --- | --- |
| 0 or 1 session | All participants considered as not responding to HAP | Not relevant | It would not be possible for the intervention to elicit a therapeutic effect by the end of the first session which is why the PHQ-9 was only assessed after this time. |
| 2 – 4 sessions | Any participant with a PHQ-9 score taken at any session that was greater than 9 | Participants with all available in-session PHQ-9 scores less than 10 | If a participant had a PHQ score greater than 9, it is assumed they dropped out of the trial as they did not feel they were responding to treatment.  Likewise, if a participant had a PHQ-9 score less than 10 it is assumed they were feeling better and stopped attending sessions. |
| 5 - 8 sessions | Any participant with a PHQ-9 score taken in-sessions 3-6 that was greater than 9 | Participants with all available PHQ-9 scores less than 10 at sessions 3-6 | A participant who had a PHQ-9 score greater than 9 at the second session, but PHQ-9 scores less than 10 at sessions 3-6, were likely responding to treatment.  A participant who had a PHQ-9 score less than 9 at sessions 3-4, but greater than 9 at sessions 5-6 were likely not responding and/or at risk of suicide. |

Estimation methods

Estimation for the different effects was based on Monte Carlo integration using 1,000 fold expanded dataset. (15) Estimates for the total, direct, and indirect effects were obtained by running regression models for the outcome of PHQ-9 score, separately in the exposed and unexposed, setting the mediators at a random subject-specific distribution (Table 1).

To fix the mediators to a random subject-specific distribution for both the unexposed/exposed levels for each participant, predictive regression models were run using the expanded dataset. Specifically, the following regression models were used: linear regression models were used for M1a and M2; ordered logistic regression models were used for M1b and M3b; logistic regression models were used for M3a.

The unexposed status for mediators measured in both arms of the trial were set using data from participants in the control arm. However, this was not possible for mediators that were part of the HAP intervention (i.e. characteristics of the sessions) and therefore not measured in the control arm. In these instances, the unexposed status was set to zero (i.e. representing no sessions were attended as was the case for a small number of participants in the experimental arm).

Normality assumptions for all linear regression models were evaluated by examining the residual plots after running the regression commands using the post-estimation command *predict r, resid* followed by the *pnorm* and *qnorm* commands. Bias-corrected confidence intervals were based on nonparametric bootstrap with 1,000 resamples.(15) The bootstrap also accounted for clustering at the primary health clinic.
