## Appendix 2 for "Causal meditation analysis to understand how different components of a complex psychological intervention improved symptoms (or not) of depression in Goa, India"

**Appendix 2: Missing data analyses**

Data was imputed separately in the experimental and control arms. Imputation models also included predictors of missingness. The imputation model for missing PHQ-9 scores at 12 months included the following predictors: baseline PHQ-9 scores, age, education, duration of depression before enrolling in the trial, expectations of the trial, marital status, behavioural activation scores at three months, proportion of homework completed, number of sessions attended, non-response to the intervention and the number of extra sessions attended. The imputation model for missing behavioural activation scores at 3 months included the following predictors: baseline PHQ-9 scores, age, education, duration of illness prior to enrolling in the trial, proportion of homework completed, number of sessions attended, non-response to the intervention and the number of extra sessions attended. The data was also imputed prior to the Monte Carlo integration procedure. The imputation step was repeated on each bootstrap sample.

Results

All findings suggest that the imputed data are largely similar to the complete data.

**Table 1: Comparison between primary and secondary outcomes, mediator-outcome confounders for complete and imputed data between the experimental and control arm for complete cases and imputed data**

| **Variable** | **Control,**  **N (%)** | | **Experimental,**  **N (%)** | |
| --- | --- | --- | --- | --- |
|  | Complete case  N=229 | Imputed data  n=248 | Complete case  n=218 | Imputed data  n=245 |
| **Remission at 12 months (PHQ-9 <10)** | 107 (46.7) | 118 (47.6) | 137 (62.8) | 154 (62.9) |
| **Mean PHQ-9 scores at 12 months (SD)** | 10.5 (7.4) | 10.5 (7.7) | 8.2 (6.9) | 8.1 (6.9) |
| **Mean activity levels measured using the adapted BADS-SF at three months (SD)** | 9.8 (4.3) | 9.8 (4.4) | 12.0 (4.7) | 12.1 (4.8) |

^a^ Missing data has been imputed by trial arm, using single imputation stochastic models adjusted for any factors that could potentially influence missingness

Table 2 describes differences between mediators of interest between remission from depression and no remission from depression, for imputed and complete cases separately.

**Table 2: Comparison of mediators between participants with and without remission from depression at 12 months, in the experimental arm of the trial for complete cases and imputed data**

| **Mediators** | **Complete case n=218** | | **Imputed data n=245** | |
| --- | --- | --- | --- | --- |
|  | **No remission from depression (N=81)**  **(PHQ-9> 10)**  **N (%)** | **Remission from depression (N=137)**  **(PHQ-9< 10)**  **N (%)** | **No remission from depression (N=89)**  **(PHQ-9> 10)**  **N (%)** | **Remission from depression (N=156)**  **(PHQ-9< 10)**  **N (%)** |
| Number of sessions attended  0  1  2  3  4  5  6  7  8 | 3 (3.7)  2 (2.5)  6 (7.5)  1 (1.2)  1 (1.2)  11 (13.6)  22 (27.2)  10 (12.4)  25 (30.9) | 10 (7.3)  8 (5.8)  14 (10.2)  2 (1.5)  1 (0.7)  46 (33.6)  31 (22.6)  9 (6.6)  16 (11.7) | 6 (6.7)  4 (4.4)  7 (7.8)  2 (2.2)  2(2.2)  11 (12.2)  23 (25.6)  10 (11.1)  25 (27.8) | 13 (8.4)  16 (10.9)  17 (11.0)  3 (1.9)  2 (1.3)  47 (30.3)  31 (20.0)  10 (6.5)  16 (10.3) |
| **M1a: Number of phases of sessions completed**  None (no sessions completed)  One (sessions 1 to 2 completed)  Two (sessions 3 to 4 completed)  Three (sessions 5 to 8 completed) | 3 (3.7)  8 (9.9)  2 (2.5)  68 (84.0) | 10 (7.3)  22 (16.1)  3 (2.2)  102 (74.5) | 6 (6.7)  11 (12.2)  4 (4.4)  69 (76.7) | 13 (8.4)  33 (21.3)  5 (3.2)  104 (61.1) |
| **M1b: Proportion of assigned activities completed**  No homework completed  1 – 50% homework completed  50 – 100% homework completed | 129 (63.6)  13 (6.4)  61 (30.1) | 128 (52.5)  22 (9.0)  94 (38.5) | 13 (14.4)  14 (15.6)  63 (70.0) | 34 (21.9)  25 (16.1)  96 (61.9) |
| **M2: Adapted BADS – SF score (mean, standard deviation) measured at 3 months** | 10 (4.5) | 13.6 (4.1) | 9.5 (4.8) | 13.4 (4.3) |
| **M3a: Participants response to HAP measured by PHQ-9 score greater than 9 at 3^rd^ or 4^th^ session**  Did not respond to HAP  Responded to HAP | 48 (59.3)  33 (40.7) | 47 (34.3)  90 (65.7) | 56 (62.2)  34 (37.8) | 63 (40.7)  92 (59.4) |
| **M3b: Number of extra sessions attended if participant did not respond to HAP**  None  One  Two | 46 (56.8)  10 (12.4)  25 (30.9) | 112 (81.8)  9 (6.6)  16 ((11.7) | 55 (61.1)  10 (11.1)  25 (27.8) | 129 (83.2)  10 (6.5)  16 (10.3) |

^a^ Missing data has been imputed by trial arm, using single imputation stochastic models adjusted for any factors that could potentially influence missingness

**Table 4: Total effect and interventional in(direct) effects for the HAP intervention at 12 months using complete data only**

|  | **Estimates (bias-corrected 95% CI)^a^** | |
| --- | --- | --- |
| **Effect** | **Complete case** | **Imputed data** |
| Total effect | -2.3 (-3.1 to -1.2) | -2.2 (-3.3 to -0.8) |
| Interventional direct effect | -3.0 (-6.2 to 0.5) | -3.2 (-5.6 to -0.5) |
| Interventional indirect effect through sessions only | 2.6 (-2.5 to 5.3) | 2.9 (-1.3 to 5.0) |
| Interventional indirect effect through BA levels only | -1.0 (-1.3 to -0.6) | -1.0 (-1.3 to -0.6) |
| Interventional indirect effect through extra sessions | -0.8 (-2.4 to 1.5) | -0.8 (-2.1 to 1.2) |
| Interventional indirect effect through the dependence of BA levels on sessions | -0.1 (-0.9 to 0.4) | 0.0 (-0.6 to 0.8) |

^a^ Estimates have been adjusted for mediator-outcome confounders of baseline PHQ-9 scores, age and education.

^b^ Estimation for the different effects was based on Monte Carlo integration using 1,000-fold expanded dataset

^c^ Bias-corrected confidence intervals were based on nonparametric bootstrap with 1,000 resamples

^d^ Missing data has been imputed by trial arm, using single imputation stochastic models adjusted for any factors that could potentially influence missingness
